# Using facial reaction analysis and machine learning to objectively assess palatability of medicines in children

**DOI:** 10.1101/2023.08.21.23293473

**Authors:** Rabia Aziza, Elisa Alessandrini, Clare Matthews, Sejal Ranmal, Ziyu Zhou, Elin Haf Davies, Catherine Tuleu

## Abstract

For orally administered drugs, palatability is key in ensuring patient acceptability and treatment compliance. Therefore, understanding children’s taste sensitivity and preferences can support formulators in making paediatric medicines more acceptable. Presently, we explore if the application of computer-vision techniques to videos of children’s reaction to gustatory taste strips can provide an objective assessment of palatability.

Primary school children tasted four different flavoured strips: no taste, bitter, sweet and sour. Data was collected at home, under the supervision of a guardian, with responses recorded using the Aparito Atom5™ app and smartphone camera. Participants scored each strip on a 5-point hedonic scale. Facial landmarks were identified in the videos, and quantitative measures such as changes around the eyes, nose and mouth were extracted to train models to classify strip taste and score. We received 197 videos and 256 self-reported scores from 64 participants. The hedonic scale elicited expected results: children like sweetness, dislike bitterness and have varying opinions for sourness. The findings revealed the complexity and variability of facial reactions and highlighted specific measures, such as eyebrow and mouth corner elevations, as significant indicators of palatability. This study into children’s taste specificities can improve the measurement of paediatric medicine acceptability. An objective measure of how children feel about the taste of medicines has great potential in helping find the most palatable formulation. Moreover, collecting data in the home setting allows for natural behaviour, with minimal burden for participants.

**Author summary:** When formulating medicines for children, understanding the taste profile is crucial to ensure they are not excessively unpleasant. In this study, we assessed if facial reactions in response to taste stimuli can be used to easily measure children’s feelings about the taste. We recorded videos of children trying different taste strips and analysed their facial expressions in response to each taste. The strips had different flavours: bitter, sweet, sour, and one with no taste. We also asked the children to rate each strip on scale of 1 to 5. We collected data from 64 children. The results confirmed that children generally like sweet tastes and do not like bitter ones. Their opinions on sour taste varied. Moreover, we found that specific facial reactions, like changes in their eyebrows and mouth corners were good indicators of taste preferences. The analysis of facial expressions can help formulators make medicines for children taste better. By objectively measuring how children feel about the taste of medicines, we can create more acceptable medicines for them. The collection of this data at home ensured children were in their comfortable environment, making it easier for them to be part of the study.

## Introduction

The concept of patient acceptability is gaining progressive relevance in the development of paediatric dosage forms. Acceptability is defined as the overall ability of the patient and caregiver to use a medicinal product as intended or authorised and it is determined by characteristics of the user (age, ability, disease type) and of a medicinal product (e.g. palatability, swallowability, appearance) (1).

Thus, acceptability can have a significant impact on the patient’s adherence and consequently on the safety and efficacy of a product. For this reason, the European Medicines Agency (EMA) has repeatedly emphasised the importance and incentive to assess the acceptability of formulations for paediatric use, including in its 2006 Reflection Paper (2) and 2014 guideline on pharmaceutical development of medicinal products for children, where it is stated that the evaluation of acceptability should be embedded in the development program and evaluated within the targeted population (1). Consequently, in recent years, there has been an increased emphasis on conducting studies examining factors affecting medicines acceptability in children.

For orally administered drugs, palatability is key in determining patient acceptability and treatment compliance (1). Palatability is defined as the overall appreciation of a medicinal product in relation to its smell, taste, aftertaste and texture (i.e. feeling in the mouth) (1). Specifically, taste is frequently reported to be a common reason for non-compliance among children (3). Thus, regulatory agencies strongly encourage the inclusion of acceptability testing, including palatability assessment, as part of the product development and clinical program in the target patient population (3).

Several methodologies for palatability assessment in children are available, and they have been largely reviewed (3–7). These methodologies should be age-appropriate and depending on the age of the child may involve collecting data from patients and/or caregivers (4). The selection of the appropriate taste assessment methodology is determined by the cognitive capacity of the child, and until now, there is no methodology validated for accuracy and reliability with any particular age group (3,6), although the facial hedonic scale is considered the gold standard for drug palatability testing in children (8). However, this scale cannot be used in very young children or in those with communication issues, cognitive impairments, and/or developmental delay.

The EMA reflection paper defines four key criteria for palatability assessment in children: 1) the test should be short, 2) the test has to be intrinsically motivating and “fun” to do, 3) the procedure should be as easy as possible, 4) the number of variants to be tested should be limited to a maximum of four. However, the reflection paper does not aim to provide any scientific, technical, or regulatory guidance (2). This suggests the opportunity for the development of more objective quantitative technological advancements such as the use of high throughput systems [https://www.opertechbio.com/technology], facial electromyography (9), electrogustometry (10), or facial recognition technology (8,11) in palatability assessment (3).

Observation of children’s facial expressions after a taste stimulus to assess taste reactivity, is not new. Some of the earliest investigations on taste in children consisted of videotaping infants and then characterising their oromotor reflexes when taste stimuli were placed on the tongue or in the oral cavity (12–15). In 1988, Oster and Rosenstein (15) developed a method for describing orofacial responses by using the Ekman and Friesen’s (16) anatomically based Facial Action Coding System (FACS). FACS virtually decomposes any facial expression into its constituent action units (AUs). Video records are analysed in slow motion to quantify the actual number of affective reactions infants express to a taste stimulus, as a measure of valence and intensity (7,17). The advantage of using this method is that it can be used in non-verbal children such as infants. However, the analysis of video images requires trained individuals to establish reliability across scores and it can be subjective (18). Moreover, this method is time-consuming and costly (7).

A large number of studies have focused their attention on the use of artificial intelligence (AI) technologies, i.e. facial emotion recognition technologies, to assess food and drinks preferences and consumer acceptance (19–25) with promising results. However, only recently, the use of these technologies has caught the attention of researchers in the assessment of taste responses to medicinal products.

Kearns et al. (26) undertook a prospective, pilot study to assess the feasibility of using facial recognition technology to assess drug palatability in 10 children aged between 7 to 17 years. Although the qualitative assessment of the facial recognition data demonstrated the ability to discriminate between bitter and sweet tastants, their facial recognition software (Noldus FaceReader® 7) and approach showed some limitations in discriminating taste profiles and highlighted that further refinement was required before this methodology can be applied more widely (8). The Noldus FaceReader® 7 software measures the intensities of a predefined set of emotions e.g., happy, angry, disgusted etc., on a frame-by-frame basis.

Similarly, Peng et al.(27), used the same software to compare the palatability of two preparations of carbocysteine among healthy children aged 2-12 years. The palatability assessed by emotional valences was performed using a facial action coding system by FaceReader™, which reflected the quantification of emotions; a positive value represents a positive emotion, and a negative value represents a negative emotion (27).

Presently, we refine the work of Kearn’s group (26), to explore if the application of computer-vision techniques to videos of primary school children’s reaction to gustatory taste strips can provide an objective assessment of palatability. Our methodology uses pose estimation for facial recognition analysis, which allows access to the raw movements of facial features, rather than through the lens of emotional reactions. Finally, our study was conducted in a domiciliary setting to allow for natural behaviour, with minimal burden for participants.

## Materials and Methods

### Participants

Participants were children aged between 4 to 11 years old recruited from a primary school in London, United Kingdom, and their adult caregiver. Prior to the study, all participants received an information sheet with the study details. Participants and their caregivers had to sign informed consent and assent forms respectively if they were willing to participate in the study. The study was approved by the UCL research ethics committee (REC) 4612/029.

As this was an exploratory study, no formal requirements were put on sample size. All pupils at a school of 840 were invited to join the study.

### Study Design and Procedures

This study was a single blind taste assessment, conducted in a home environment. After consent, all participants were provided with a study pack for the taste evaluation to be completed at home. The pack contained the study instructions, four taste strips and information about how to download the Aparito app (Atom5^TM^) on their smartphone. Atom5^TM^ is a secure, encrypted and password protected platform (ISO 13845, ISO 27001, Cyber Essential Plus, CREST tested and ePrivacyApp awarded by ePrivacyseal GmbH) for remote monitoring of a wide range of adult and paediatric disease areas, designed to collect digital endpoint data. Commercially available Burghart (or ODOFIN) taste strips (MediSense, Groningen, The Netherlands) were used in this study. The taste strips are made of filter paper impregnated with different solutions containing substances found in food and drinks. They are used in clinical and research contexts as a validated procedure to investigate taste ability/gustatory sensitivity in both children and adults (28). One of the strips was blank with no tastant, one strip was bitter (0.006 g/mL of quinine hydrochloride), one was sweet (0.4 g/mL of sucrose), and one sour (0.3 g/mL of citric acid). Each taste strip was individually repackaged into coloured Mylar foil bags for blinding purposes: blank in a white coloured foil bag, bitter in yellow, sweet in green and sour in purple.

After receiving the study pack, participants were asked to input their age and sex on the app. Then, instructions guided the participants through each step of the study. Children were instructed to place one strip on the middle of their tongue, close their mouth and test the sample for 10 seconds before removing the strip. The adult caregiver used their smartphone video to record the facial reaction of the child as they tasted the strip. After removing each strip, the children were asked to rate the sample on a 5-point hedonic smiley face scale, where 1 corresponded to a sad face, indicative of dislike and 5 to a happy face indicating the liking of the taste of the strip, Fig 1. Finally, children were invited to provide their feedback about the tasting experience through an open response question.

**Fig 1.**
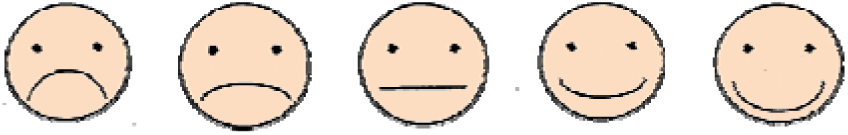
A 5-point hedonic smiley face scale that was used in the study.

Instructions indicated to test the blank control strip first so that participants could practise the correct use of the strip, the video recording, and how to record their responses in the app. The other three taste strips were tested in three different sequences as indicated by the foil bag colour (sweet, sour, bitter - sour, bitter, sweet - bitter, sweet, sour) and children were randomly assigned to one of the three sequences by the app. To avoid any anticipation and bias in their responses, participants were blinded to the taste of the strip they were tasting and were instructed to taste each strip sequentially from each coloured foil bag. Children were invited to take some water to help remove any residual taste between each sample.

All data recorded within the app was transferred onto the Atom5 ^TM^ software platform and stored securely on Microsoft Azure. All data were collected and stored in accordance with the Data Protection Legislation 2018 and General Data Protection Regulation (GDPR) 2018.

### Statistical Analysis

For each taste strip, the rank rating of the hedonic scale was analysed to see if any difference could be appraised between different age groups, sex, and order of strip testing. Also, differences in rank rating between different tastes were assessed. The Kruskal-Wallis H test was used for the analysis and the IBM® SPSS Statistics software platform (IBM Corp. Released 2021. IBM SPSS Statistics for Windows, Version 28.0. Armonk, NY: IBM Corp) was used with the limit of statistical significance set at α = 0.05.

### Machine Learning for Pose Estimation

### Pose Estimation Framework

We used a CNN (Convolutional Neural Network) based, open-source ML (Machine Learning) framework, Mediapipe (29). Mediapipe is a perception pipeline builder that offers different pose estimation components, including face detection, face mesh, hands, body, and object tracking. In this study, we used the face mesh component to estimate 368 3D face landmarks per frame.

### Data Pre-processing

The video data were filtered on two levels: frame level and facial landmark level.

First, we identified which frames to include in the analysis. We defined the tasting task as the sequence of frames per video from when the taste strip was inserted in the subject’s mouth, until just before the action of removing said strip. The cleaning process included going through the videos and manually identifying the beginning and end times of the tasting task. Importantly, we noticed that some subjects reacted after the taste strip was removed from the mouth. Therefore, we also identified a post-tasting section that started from the moment after the strip was removed from the mouth, until the end of the video or until the subject was distracted by something else (e.g., if the child starts talking to someone else in the room or turns away and moves out of the frame).

Secondly, we identified 53 facial landmarks to be included in the analysis out of the 368 landmarks extracted by Mediapipe, which allows for the outlining of the main visible facial features, Fig 2.

**Fig 2.**
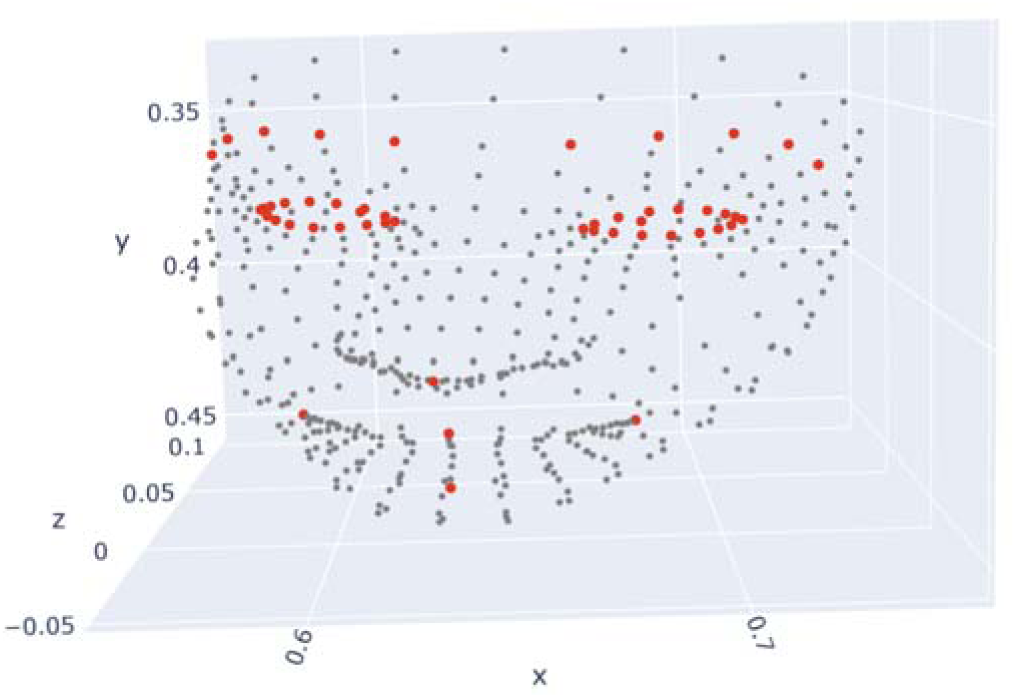
Landmarks identified to be included in the analysis (red) plotted against all landmarks estimated by Mediapipe (grey).

### Facial Feature Aggregation

The extracted landmarks were aggregated into six measures inspired from Novotny et al., 2022 (30):

- Eyebrow elevation: left and right variants of the distance between the median of the eyebrow landmarks and the nose tip landmark.
- Eyebrow tilt: left and right variants of the angle between the fitted line to the five eyebrow landmarks and the line connecting the inner corners of the eyes.
- Eyebrow shape: left and right variants of the angle between the lines connecting the middle eyebrow landmark with the left and right endpoints of the eyebrow.
- Palpebral aperture: left and right variants of the surface delimiting the eye area.
- Lip elevation: upper and lower variants of the distance between the middle lip landmark and the nose tip.
- Mouth corner: left and right variants of the distance between the mouth corner landmark and the nose tip.

All distance measures were rescaled by a reference measure to account for the variations in the landmarks’ coordinates caused by something other than the facial reaction. These other variations were caused by the subject’s face changing position or angle during the video, e.g., head rotation, head movement closer or farther from the camera, the camera’s angle moving during the test, etc. The reference measure chosen was the distance between the right inner corner of the eye and the tip of the nose.

## Results

### Data Description and Participants’ Demographics

A total of 250 participants agreed to take part in the study and received a study pack at home. Of these, 40 participants completed all the hedonic ratings and video recordings, and 24 participants completed all the hedonic scales but did not record one or more videos. Thus, the data analysis was performed with data from 64 participants; Table 1 reports the number of hedonic scales and videos recorded per taste strip.

**Table 1.** Total number of hedonic scale ratings and videos analysed per each taste.

| Taste of the strip | No. of hedonic scales | No. of videos |
| --- | --- | --- |
| Neutral | 64 | 51 |
| Sour | 64 | 46 |
| Sweet | 64 | 54 |
| Bitter | 64 | 46 |
| Total | 256 | 197 |

Children in the study were aged between 5 to 11 years, with a median age of 8.5 years (SD 1.46), Table 2, and there were slightly more females (n=36) than males (n=28).

**Table 2.** Age distribution of the children participating in the study.

| Age (years) | Frequency (%) |
| --- | --- |
| 5 | 1 (2%) |
| 6 | 3 (5%) |
| 7 | 10 (16%) |
| 8 | 18 (28%) |
| 9 | 13 (20%) |
| 10 | 11 (17%) |
| 11 | 8 (13%) |

### Hedonic Scale Results

The ratings of the hedonic scale for each taste strip were analysed by gender, age, and randomisation order of the strips to assess if any difference between groups existed. The analysis by gender showed that there were no significant differences between boys and girls in terms of hedonic responses to the taste of each strip (_χ_2(2) = 0.46, p = 0.50 for the blank (control) strip, _χ_2(2) = 1.50, p = 0.23 for the sour strip, _χ_2(2) = 2.32, p = 0.13 for the sweet strip, and _χ_2(2) = 2.30, p = 0.13 for the bitter strip). Different ages also showed similar ranking scores for each taste strip (_χ_2(2) = 2.04, p = 0.92 for the control strip, _χ_2(2) = 4.19, p =0.65 for the sour strip, _χ_2(2) = 3.92, p =0.69 for the sweet strip, _χ_2(2) = 1.99, p =0.92 for the bitter strip). Similarly, changing the order of how the taste strips were assessed did not alter the rating of each taste strip (_χ_2(2) = 0.64, p = 0.73 for the sour strip, _χ_2(2) = 0.41, p =0.82 for the sweet strip, _χ_2(2) = 0.80, p =0.67 for the bitter strip).

Instead, a statistically significant difference in ratings emerged across the different tastes (_χ_2()= 124.62, and p= 0.001), with the following predominant scores observed for each taste strip: 3 for the blank control strip, 1 for the bitter strip, 5 for the sweet strip, and various scores for the sour strip. These results indicate that the hedonic scale elicited expected results: children like sweetness, dislike bitterness and have varying opinions for sourness, Fig 3.

**Fig 3.**
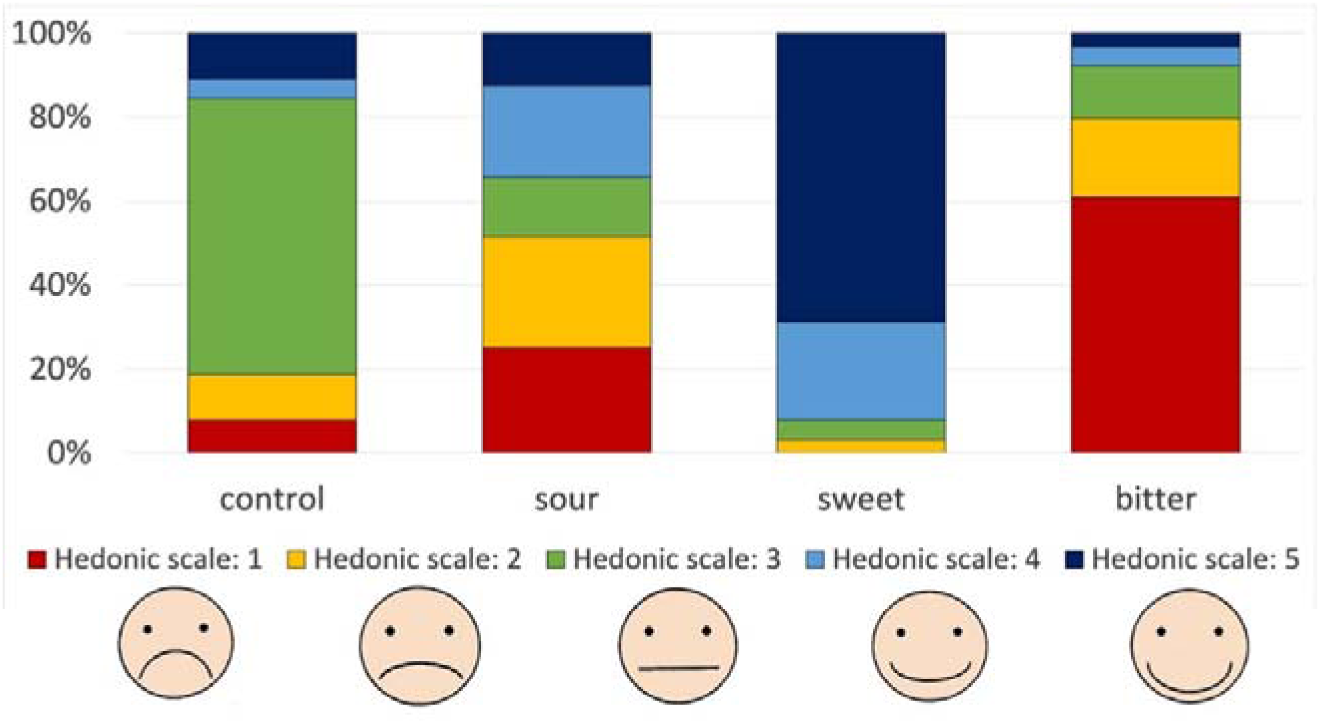
Participants’ hedonic scale rating for each taste strip, where 1 corresponds to not liking the taste (sad face), and 5 corresponds to liking the taste of the strip (happy face).

### Facial Measures

Facial landmark coordinates were extracted for each frame captured during or after the tasting task. A total of 97.4% of frames (accounting for 95561 total frames) were successfully processed using the Mediapipe framework. Failures to process some frames were due to subjects turning away from the camera or covering their face. Fig 4 shows the distribution of processed frames during and after the tasting.

**Fig 4.**
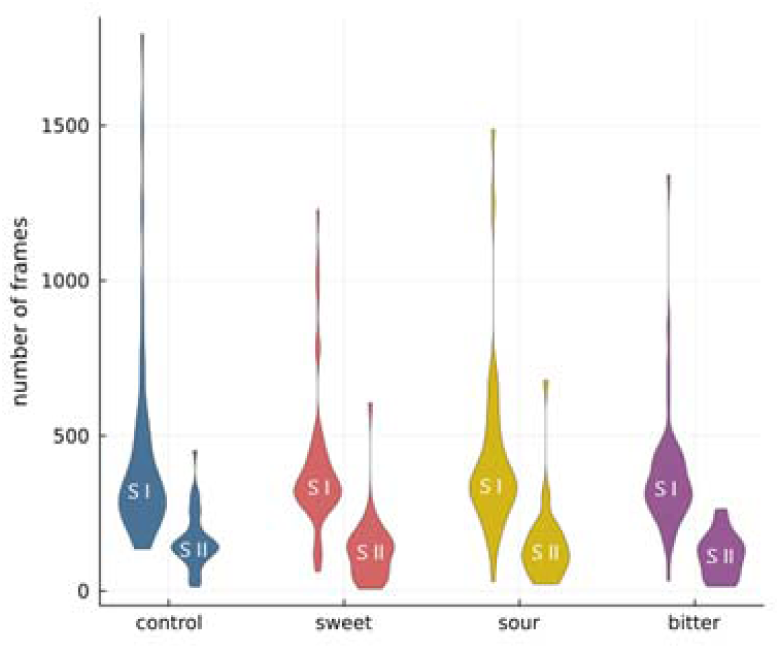
Number of frames processed with Mediapipe per taste and section: during the tasting (SI) and post tasting (SII).

Using the extracted coordinates, we computed the facial measures (described in section 2.4.3). A sample data of two subjects (four videos per subject: one for each taste) is depicted in Fig 5 as a 5-frame moving average of three

**Fig 5.**
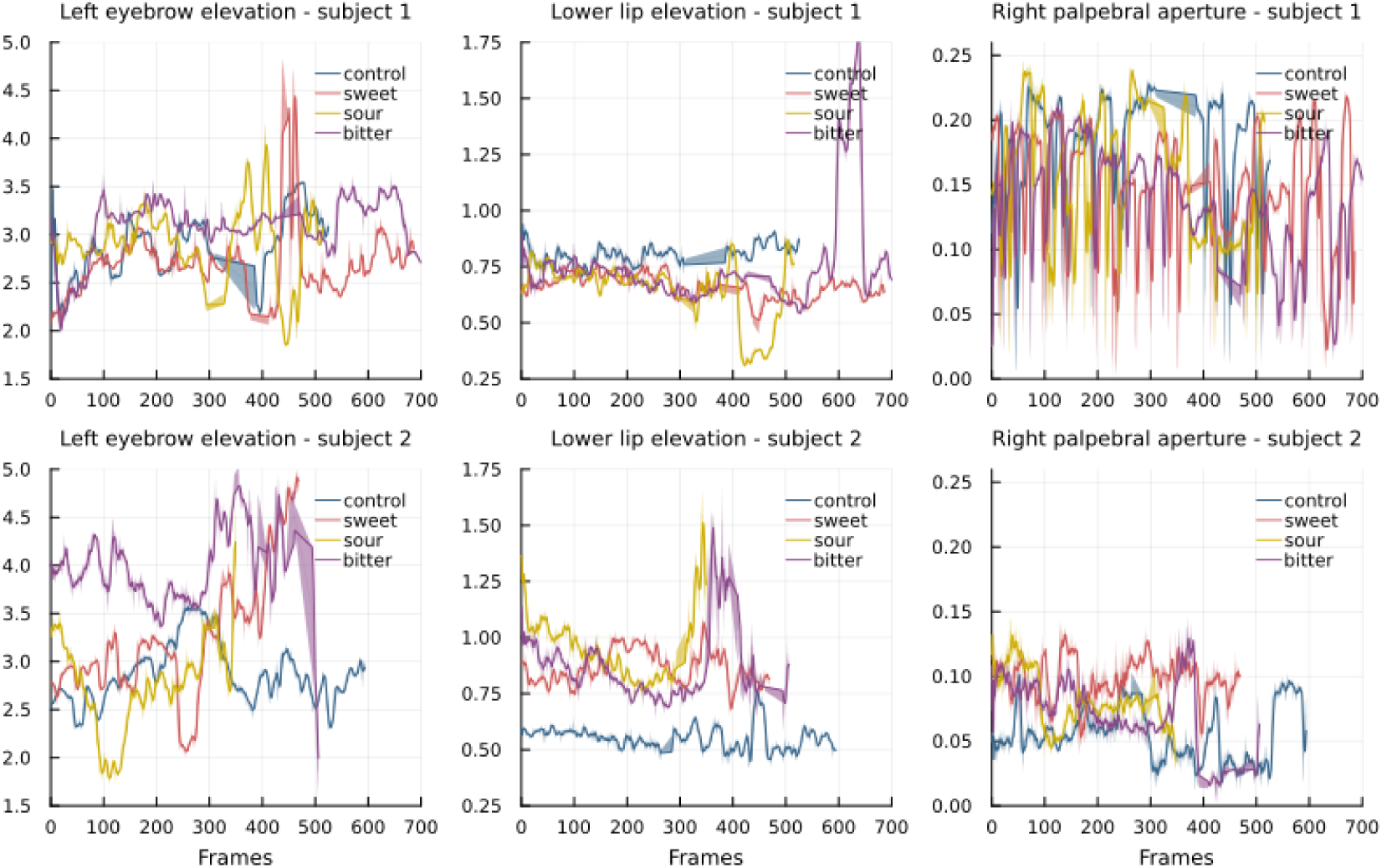
Sample of rescaled measures (left eyebrow elevation, lower lip elevation, right palpebral aperture) of two subjects, representing the variation of the measures over time per tasting (including both during and post tasting sections).

We then calculated the standard deviation of the rescaled measures for all subjects, including all their videos that passed quality checks. We plotted this, grouped per taste and hedonic score, in Figs 6 and 7.

**Fig 6.**
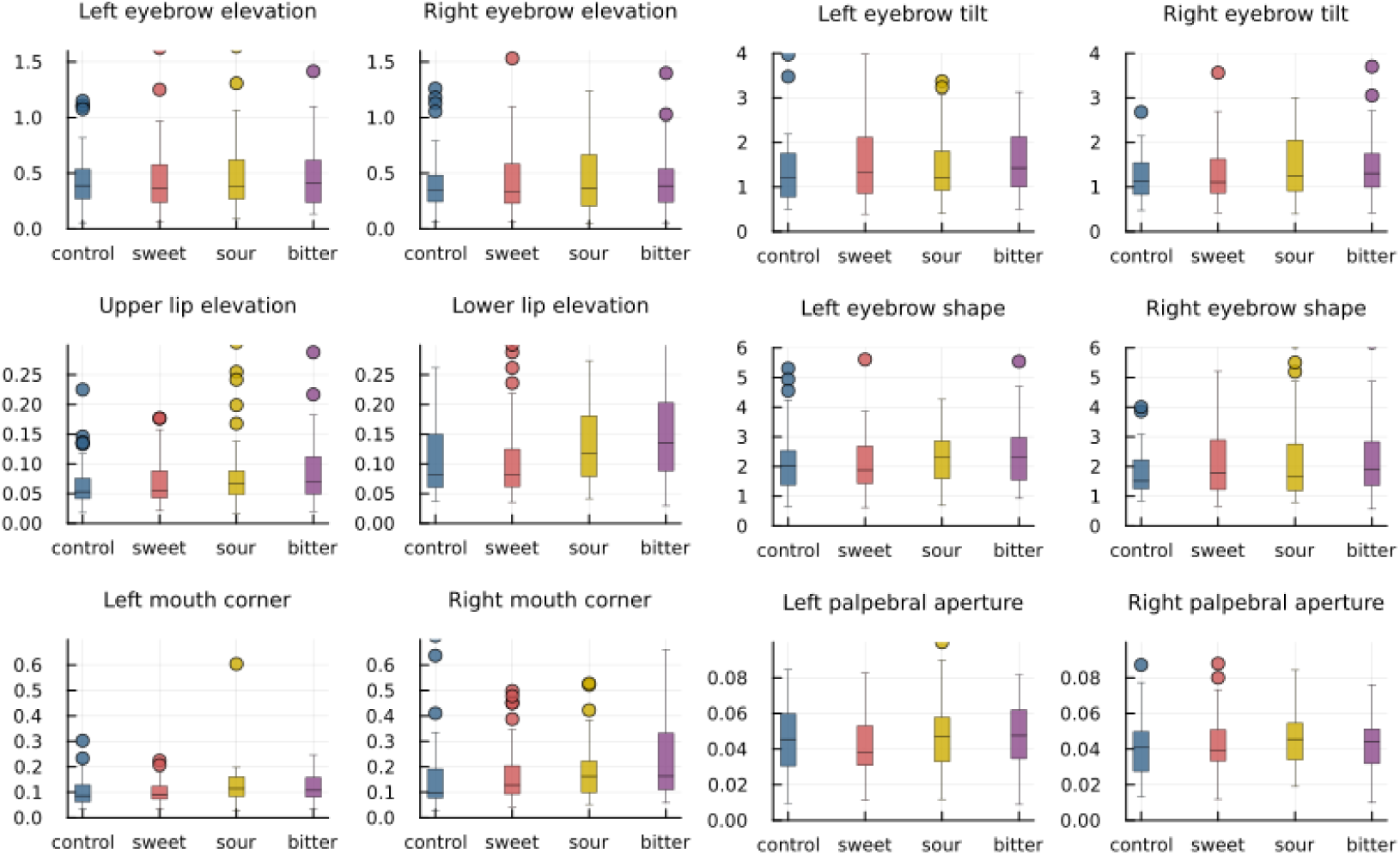
Box plots of the standard deviation of the rescaled measures, grouped by taste.

**Fig 7.**
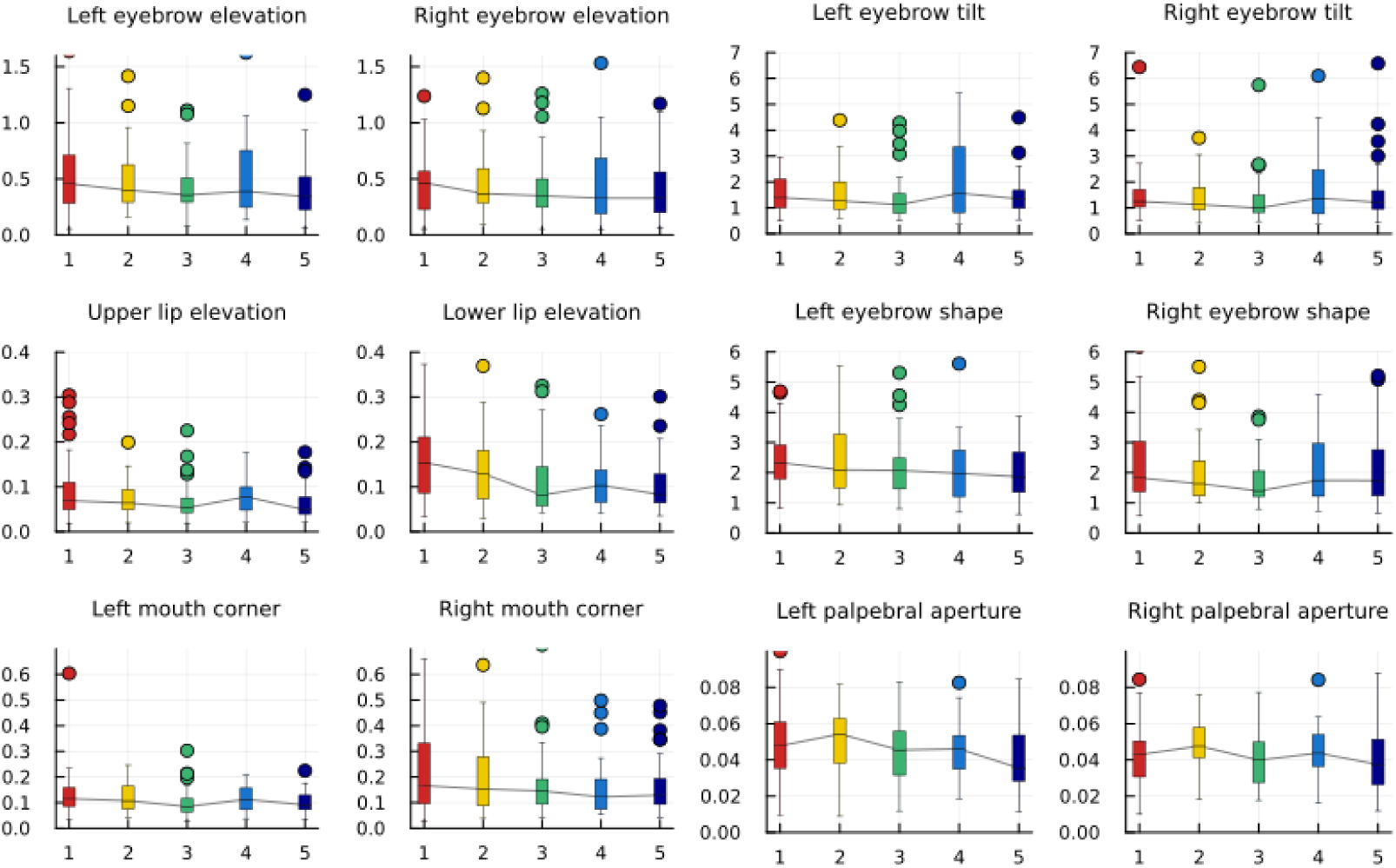
Box plots of the standard deviation of the rescaled measures, grouped by hedonic scores.

Moreover, we ran an analysis that determines which facial measure(s) were most predictive of palatability. We used an Extra-trees classifier to rank the importance of each measure. The “Right eyebrow elevation” measure was found to be the most important, followed by the “Left eyebrow elevation” and the “Left/Right” mouth corner”, Fig 8.

**Fig 8.**
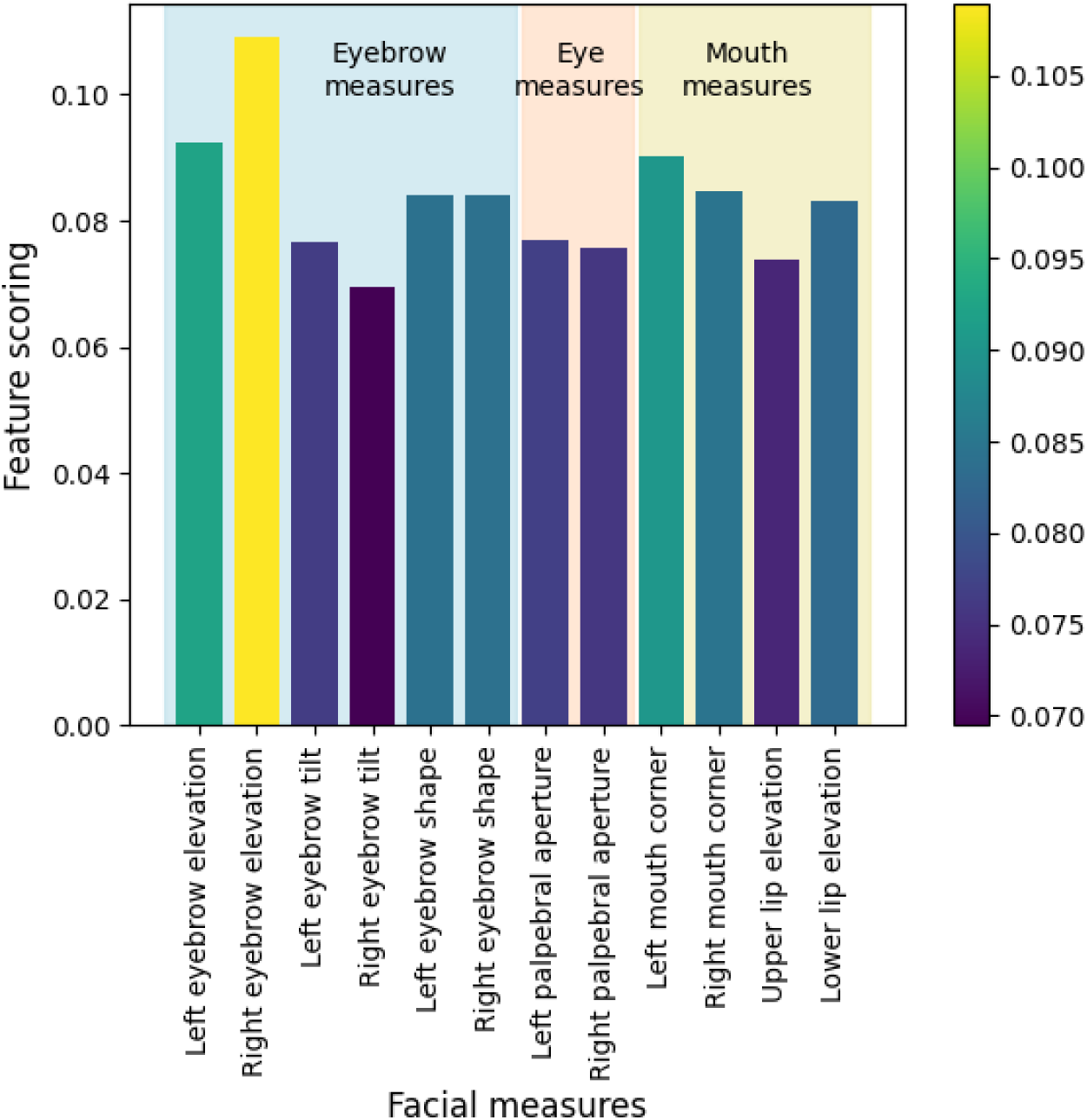
Ranking of the importance of the facial measures using an Extra-Trees classifier.

## Discussion

The present study sought to investigate facial reactions to different tastes in children, with the aim of identifying key indicators of palatability perceptions.

Two previous studies have assessed the use of facial recognition technology to evaluate palatability in the context of medicinal products in children (26,27). However, the main differences of our study compared to the previous studies are as follows. Firstly, our study was conducted in a domiciliary setting rather than in a standardised laboratory environment. If, on the one side, this meant reduced video quality and compliance with the instructions, on the other side, our study showed that it is feasible with the advantage of posing a minimal burden for participants.

Secondly, the methodology applied in our study differed from that of Kearn’s and Peng’s study (26,27). We applied pose estimation for facial reaction analysis assessing the raw movements of facial features, rather than classifying stimuli without the need to translate them into emotions which can be biased by aspects other than the taste.

Finally, taste strips were used instead of liquids to measure the palatability. The advantage of using taste strips rather than solutions is that the latter pose inherent quality and safety related challenges to taste testing, given their bulkiness and thus difficulty of storage and transport, as well as their swallowing risk, particularly if used by children (31).

This study showed that taste strips were easy to use and results from the hedonic scale showed that expected results were elicited: children like sweetness, dislike bitterness and have varying opinions for sourness. Previous studies have reported that children’s liking of sweet and dislike of bitter reflect their basic biology (32). The appreciation for sourness seems to be influenced by children’s food habits and there are various experiential factors that can influence flavour preferences during childhood (33,34).

In the 197 videos available, the relative proportions of frames per taste were as follows: 28.29% control, 26.15% sweet, 24.40% sour, and 21.16% bitter. The lower value for bitter was due to less adherence to the 10 seconds minimal duration of the video.

This well-balanced distribution of processed frames across the different tastes enabled a comprehensive evaluation of facial reactions.

Overall, our findings demonstrate a clear signal of a reaction to taste, but also highlight the complexity and variability of facial reactions in response to different tastes.

For instance, Figure 5 illustrates how the bitter taste resulted in a higher lower lip elevation, while the sweet taste elicited a higher left eyebrow in both subjects. However, these signals were not consistent across all measures and subjects, indicating the complex nature of taste perception.

Our study considered a range of measures, and we found that right eyebrow elevation, left eyebrow elevation, and left mouth corner were the three most significant indicators of children’s palatability perceptions, ranked in decreasing order of importance. Furthermore, we observed that some measures, such as lower lip elevation, showed a decreasing signal as the hedonic score increased, and that most measures exhibited symmetry between their left and right variants.

The results from the hedonic scale ratings support this methodology as the current gold standard for use with this age and ability group. However, further insight may be possible by analysing the facial reactions in relation to the hedonic ratings, for example, considering separately the reactions of those who liked and disliked the sour taste.

One limitation of this study is that the sample size obtained was smaller than what initially expected, although slightly larger than that of Kearn’s and Peng’s (26,27). This can partly be explained by the fact that the study required proactive commitment from the caregiver’s side and the fact that several parents were not willing to provide videorecords of the face of their children, although clear explanation about privacy and data confidentiality were provided in the Information Sheet.

A second limitation of this study was that only selected facial expression was considered. To generalise the findings, larger sample size and more diverse facial expression should be examined in future studies.

The results of this study provide new insights into the dynamics of facial expressions in response to taste stimuli. The study generated meaningful results, despite the relative lack of consistency and standardisation inherent in data gathered from a home setting, supporting the potential use of a decentralised approach.

## Conclusion

Our study provides valuable insights into the complex nature of taste perception in children and their potential application to drug delivery by improving the acceptance of orally administered medications in children. Further investigations could be conducted to explore other non-verbal cues to provide a more comprehensive understanding of the factors that influence taste perception in children. For instance, voice patterns may reveal vocal cues that indicate preferences or aversions. Besides that, body movement and hand gesture analysis can also offer valuable information on the emotional and cognitive responses to taste stimuli. These findings may be used to develop interventions that enhance the understanding and acceptability of medications in children, ultimately to improve their overall health outcomes.

## Data Availability

The data generated during and/or analysed during the current study are not publicly available but are available from the corresponding author on reasonable request.

## Acknowledgments

The authors gratefully thank all the children and caregivers who took part in this study. Our work would not have been possible without their help.

## References

1. European Medicines Agency (EMA). Guideline on Pharmaceutical Development of Medicines for Paediatric Use (EMA/CHMP/QWP/805880/2012 Rev. 2) [Internet]. 2012 [cited 2020 Dec 12]. Available from: https://www.ema.europa.eu/en/documents/scientific-guideline/guideline-pharmaceutical-development-medicines-paediatric-use_en.pdf

2. European Medicines Agency (EMA) Committee for Medicinal Products for Human Use (CHMP). Reflection Paper: Formulations of Choice for the Paediatric Population [Internet]. 2006 [cited 2023 May 19]. Available from: https://www.ema.europa.eu/en/documents/scientific-guideline/reflection-paper-formulations-choice-paediatric-population_en.pdf

3. Ternik R, Liu F, Bartlett JA, Khong YM, Thiam Tan DC, Dixit T, et al. Assessment of swallowability and palatability of oral dosage forms in children: Report from an M-CERSI pediatric formulation workshop. Int J Pharm [Internet]. 2018 Feb;536(2):570–81. Available from: https://linkinghub.elsevier.com/retrieve/pii/S0378517317308062

4. Kozarewicz P. Regulatory perspectives on acceptability testing of dosage forms in children. Int J Pharm [Internet]. 2014 Aug;469(2):245–8. Available from: https://linkinghub.elsevier.com/retrieve/pii/S0378517314002129

5. Davies EH, Tuleu C. Medicines for Children: A Matter of Taste. J Pediatr. 2008 Nov;153(5):599–604.e2.

6. Forestell CA, Mennella JA. The Ontogeny of Taste Perception and Preference Throughout Childhood. In: Handbook of Olfaction and Gustation. Hoboken, NJ, USA: John Wiley & Sons, Inc; 2015. p. 795–828.

7. Mennella JA, Spector AC, Reed DR, Coldwell SE. The bad taste of medicines: overview of basic research on bitter taste. Clin Ther [Internet]. 2013 Aug;35(8):1225–46. Available from: http://www.ncbi.nlm.nih.gov/pubmed/23886820

8. Abdel-Rahman SM, Bai S, Porter-Gill PA, Goode GA, Kearns GL. A Pilot Comparison of High-Versus Low-Tech Palatability Assessment Tools in Young Children. Pediatric Drugs [Internet]. 2021 Jan 25;23(1):95–104. Available from: http://link.springer.com/10.1007/s40272-020-00430-2

9. Armstrong JE, Hutchinson I, Laing DG, Jinks AL. Facial Electromyography: Responses of Children to Odor and Taste Stimuli. Chem Senses. 2007 May 17;32(6):611–21.

10. Shin IH, Park DC, Kwon C, Yeo SG. Changes in Taste Function Related to Obesity and Chronic Otitis Media With Effusion. Arch Otolaryngol Head Neck Surg. 2011 Mar 21;137(3):242.

11. Sikka K, Ahmed AA, Diaz D, Goodwin MS, Craig KD, Bartlett MS, et al. Automated Assessment of Children’s Postoperative Pain Using Computer Vision. Pediatrics [Internet]. 2015 Jul 1;136(1):e124–31. Available from: https://publications.aap.org/pediatrics/article/136/1/e124/28842/Automated-Assessment-of-Children-s-Postoperative

12. Steiner JE. Facial Expressions Of The Neonate Infant Indicating The Hedonics Of Food-Related Chemical Stimuli. In: Weiffenbach JM, editor. Taste and development: The genesis of sweet preference [Internet]. 1977. p. 173–8. Available from: https://books.google.co.uk/books?hl=it&lr=&id=P0JCgGAlwLkC&oi=fnd&pg=PA173&ots=KS2zTMxJ6D&sig=DD8HbsP0gcD4tkxYeHaj10-dl6A&redir_esc=y#v=onepage&q&f=false

13. Steiner JE, Glaser D, Hawilo ME, Berridge KC. Comparative expression of hedonic impact: affective reactions to taste by human infants and other primates. Neurosci Biobehav Rev [Internet]. 2001 Jan;25(1):53–74. Available from: https://linkinghub.elsevier.com/retrieve/pii/S0149763400000518

14. Forestell CA, Mennella JA. Early Determinants of Fruit and Vegetable Acceptance. Pediatrics. 2007 Dec 1;120(6):1247–54.

15. Rosenstein D, Oster H. Differential Facial Responses to Four Basic Tastes in Newborns. Child Dev [Internet]. 1988 Dec;59(6):1555. Available from: https://www.jstor.org/stable/1130670?origin=crossref

16. Ekman P, Wallace V. F. Facial action coding system. Environmental Psychology & Nonverbal Behavior. 1978;

17. Mennella JA, Forestell CA, Morgan LK, Beauchamp GK. Early milk feeding influences taste acceptance and liking during infancy. Am J Clin Nutr. 2009 Sep 1;90(3):780S–788S.

18. Forestell CA, Mennella JA. More than just a pretty face. The relationship between infant’s temperament, food acceptance, and mothers’ perceptions of their enjoyment of food. Appetite. 2012 Jun;58(3):1136–42.

19. Yamamoto T, Mizuta H, Ueji K. Analysis of facial expressions in response to basic taste stimuli using artificial intelligence to predict perceived hedonic ratings. Li Z, editor. PLoS One [Internet]. 2021 May 4;16(5):e0250928. Available from: 10.1371/journal.pone.0250928

20. Danner L, Sidorkina L, Joechl M, Duerrschmid K. Make a face! Implicit and explicit measurement of facial expressions elicited by orange juices using face reading technology. Food Qual Prefer [Internet]. 2014 Mar 1 [cited 2021 Oct 19];32:167–72. Available from: https://linkinghub.elsevier.com/retrieve/pii/S0950329313000086

21. Dibekliollu H, Gevers T. Automatic Estimation of Taste Liking through Facial Expression Dynamics. IEEE Trans Affect Comput. 2020;11(1):151–63.

22. Zeinstra GG, Koelen MA, Colindres D, Kok FJ, de Graaf C. Facial expressions in school-aged children are a good indicator of ‘dislikes’, but not of ‘likes.’ Food Qual Prefer [Internet]. 2009 Dec;20(8):620–4. Available from: https://linkinghub.elsevier.com/retrieve/pii/S0950329309001098

23. Zhi R, Cao L, Cao G. Asians’ Facial Responsiveness to Basic Tastes by Automated Facial Expression Analysis System. J Food Sci. 2017 Mar;82(3):794–806.

24. Zhi R, Hu X, Wang C, Liu S. Development of a direct mapping model between hedonic rating and facial responses by dynamic facial expression representation. Food Research International. 2020 Nov;137:109411.

25. Samant SS, Chapko MJ, Seo HS. Predicting consumer liking and preference based on emotional responses and sensory perception: A study with basic taste solutions. Food Research International. 2017 Oct;100:325–34.

26. Kearns GL, Bai S, Porter-gill PA, Goode GA, Farrar HC, Children A, et al. Use of facial recognition technology to assess drug palatability in pediatric patients: a pilot study. Journal of Applied Biopharmaceutics and Pharmacokinetics [Internet]. 2019;(7):37–49. Available from: https://www.scientificarray.org/wp-content/uploads/2020/02/JABPV7A5-Kearns.pdf

27. Peng Y, Zhang H, Gao L, Wang X, Peng X. Palatability Assessment of Carbocysteine Oral Solution Strawberry Taste Versus Carbocysteine Oral Solution Mint Taste: A Blinded Randomized Study. Front Pharmacol [Internet]. 2022 Feb 28;13. Available from: https://www.frontiersin.org/articles/10.3389/fphar.2022.822086/full

28. Mueller C, Kallert S, Renner B, Stiassny K, Temmel AFP, Hummel T, et al. Quantitative assessment of gustatory function in a clinical context using impregnated “taste strips”. Rhinology. 2003;44(1):2–6.

29. Lugaresi C, Tang J, Nash H, McClanahan C, Uboweja E, Hays M, et al. MediaPipe: A Framework for Building Perception Pipelines. 2019 Jun 14;

30. Novotny M, Tykalova T, Ruzickova H, Ruzicka E, Dusek P, Rusz J. Automated video-based assessment of facial bradykinesia in de-novo Parkinson’s disease. NPJ Digit Med [Internet]. 2022 Jul 18;5(1):98. Available from: https://www.nature.com/articles/s41746-022-00642-5

31. Georgopoulos D, Keeley A, Tuleu C. Assessing the feasibility of using “taste-strips” for bitterness taste panels. In: Formulating Better Medicines for Children [Internet]. Malmo; 2019 [cited 2023 May 16]. Available from: https://www.ucl.ac.uk/pharmacy/sites/pharmacy/files/eupfi_taste-strips_poster.pdf

32. Mennella JA, Bobowski NK. The sweetness and bitterness of childhood: Insights from basic research on taste preferences. Physiol Behav [Internet]. 2015 Dec;152:502–7. Available from: https://linkinghub.elsevier.com/retrieve/pii/S003193841500298X

33. Liem DG. Heightened Sour Preferences During Childhood. Chem Senses [Internet]. 2003 Feb 1;28(2):173–80. Available from: https://academic.oup.com/chemse/article-lookup/doi/10.1093/chemse/28.2.173

34. Liem DG, Mennella JA. Sweet and sour preferences during childhood: Role of early experiences. Dev Psychobiol [Internet]. 2002 Dec;41(4):388–95. Available from: https://onlinelibrary.wiley.com/doi/10.1002/dev.10067

